# High glycaemic variability in individuals with type 1 diabetes is associated with a reduced proportion of CD8^+^ TNF^+^ cells in response to influenza A virus

**DOI:** 10.1101/2024.08.27.24311465

**Authors:** Marcus Tong Zhen Wei, Katina D. Hulme, Soi Cheng Law, Ellesandra Noye, Emily S. Dorey, Keng Yih Chew, Louise C. Rowntree, Carolien E. van de Sandt, Katherine Kedzierska, Marco Goeijenbier, Katharina Ronacher, Fawaz Alzaid, Jean-Baptiste Julla, Jean-Pierre Riveline, Katie Lineburg, Corey Smith, Emma J. Grant, Stephanie Gras, Linda A. Gallo, Helen L. Barett, Kirsty R. Short

## Abstract

**Objective:** Diabetes mellitus significantly increases the risk of severe respiratory virus disease like influenza and COVID-19. Early evidence suggests that this susceptibility to respiratory viral disease is driven by glycaemic variability, rather than average blood glucose levels. In healthy individuals, blood glucose levels remain relatively stable throughout the day. However, in individuals living with diabetes, blood glucose spikes are more frequent and higher in magnitude. Continuous glucose monitoring (CGM) provides a unique opportunity to detect these hyper and hypoglycaemic events, even in the presence of an in range HbA1c.

**Research design and methods:** Here, we use blood samples and CGM data obtained from people living with Type 1 diabetes (T1D) to determine the effects of glycaemic variability on the T-cell response to influenza virus. Low glycaemic variability was defined as a coefficient of variation (CV) <33% (n = 13) whilst high glycaemic variability was defined as a CV >33% (n = 19).

**Results:** We show that high glycaemic variability in participants living with T1D is associated with a reduced proportion of CD8^+^CD107α^-^IFNγ^-^MIP1β^-^TNF^+^ T-cells in response to stimulation with influenza virus and an influenza peptide pool. High glycaemic variability in this patient population is primarily driven by hypoglycaemic events and was also associated with an increase in the proportion of naïve CD8^+^ T cells and a decrease in terminally differentiated CD8^+^ T cells (T_EMRA_).

**Conclusions:** Together, this study provides the first evidence that glycaemic variability affects the T- cell response to respiratory viruses. These data suggest that monitoring glycaemic variability may have important implications in understanding the antiviral immune response in people with diabetes.

## INTRODUCTION

Diabetes mellitus (both type 1 [T1D] and type 2 [T2D]) significantly increases the risk of severe respiratory virus infections^1-4^. The mechanisms by which this occurs remain poorly defined. Hyperglycaemia (high blood glucose levels) is typical of both T1D and T2D and its persistence is indicated by elevated glycated haemoglobin (HbA1C) levels. Hyperglycaemia is associated with increased susceptibility to severe disease, including respiratory tract infections. A meta-analysis showed that chronic hyperglycaemia (HbAa1c levels ≥7.0%) is associated with at least a 2.5 fold increased risk of pulmonary tuberculosis relative to patients with lower (<7.0%) HbA1c levels^5^. Hyperglycaemia (relative to in range blood glucose levels) also significantly increased the risk of mortality, ICU admission and intubation for COVID-19 in patients with T2D ^6^. Similarly, *in vitro* high glucose levels increase pulmonary epithelial-endothelial damage from influenza virus infection^7^. An elevated HbA1C in patients with diabetes is also associated with impaired TNF production by CD8^+^ T cells^8^.

In contrast to consistently high average blood glucose levels there is now a growing body of evidence that glycaemic variability may also play a significant role in susceptibility to severe respiratory virus infections^4,9-13^. Typically, in healthy individuals blood glucose levels remain relatively stable throughout the day except for small and short-lived post-prandial peaks. However, in individuals living with diabetes these postprandial glucose spikes are more frequent and higher in magnitude. This glycaemic variability is not indicated in measurements of a patient’s HbA1c, which largely reflects a 3-month average of blood glucose levels in terms of the percentage of glycated haemoglobin. Instead, blood glucose variability can be detected using continuous glucose monitors (CGM). Indeed, CGM data has shown that both T1D patients and T2D patients experience significant fluctuations in blood glucose levels over time^14,15^. In contrast, healthy individuals with CGM data display minimal, if any, out of range fluctuations^16^. Several studies have shown that glycaemic variability increases the severity of COVID-19^4,11-13^ and there is evidence that a similar phenomenon occurs with influenza virus^9^. However, such studies typically depend on successive measurements of an individual’s blood glucose levels (rather than CGM data) and the mechanisms underlying this association remains to be determined.

Here, we hypothesise that high glycaemic variability (as determined by CGM data) reduces the anti- viral immune response to influenza virus.

## MATERIALS AND METHODS

### Influenza virus culture

Virus stocks of HKx31 (H3N2) were prepared in embryonated chicken eggs and titres of infectious virus were determined by plaque assays on MDCK cells as previously described^17^. The use of embryonated chicken eggs was approved by the University of Queensland Animal Ethics Committee (AE000089).

### Participant recruitment

A cohort of 32 people with clinically diagnosed T1D who were routinely using continuous, or flash glucose monitoring (CGM) devices were recruited in Brisbane, Australia between 8/6/21 and 11/11/21. These individuals were a subset of a previously described cohort of 72 patients with diabetes mellitus (T1D and T2D) ^18^. Inclusion criteria were 18-60 years of age, not pregnant at the time of study, non-smokers of nicotine cigarettes, minimum diabetes mellitus duration of two years and no known immune disease requiring immunosuppressants. Blood, clinical data, and two weeks of prior CGM data from these participants were collected at the point of recruitment. To further establish the quality of CGM data, only samples with CGM time worn above 70% were included in the analysis. This study was approved by Mater Research Ethics Committee (HREC/MML/55151 V2) and the University of Queensland Ethics Committee (2019/HE002522). All methods were performed in accordance with institutional guidelines and regulations. Written consent was obtained from all study participants.

### Glycaemic variability

Glycaemic variability was determined based on an individual’s coefficient of variation (CV; glucose SD expressed as a percentage of the mean glucose) reading from their CGM with the threshold set at 33.0% as described previously^19^.

### Human PBMC isolation

10mL of whole peripheral blood was collected in BD Vacutainer® EDTA tubes (BD Biosciences). Peripheral blood mononuclear cells (PBMC) were isolated with Lymphoprep (STEMCELL, Canada) according to manufacturer’s instructions. Isolated PBMC were subsequently frozen down in Fetal Calf Serum (FCS) (Gibco) containing 10% DMSO (Sigma-Aldrich) at -80 ⍰C until analysis.

### Human T-cell phenotype characterisation

To study the effects of glycaemic variability on CD8^+^ T-cell phenotypic markers, PBMCs isolated from participants were stained as previously described^20^. Briefly, T-cells were washed and stained for lymphocyte (anti-CD3; anti-CD4 anti-CD8) and differentiation markers (anti-CD27, anti-CD45RA and anti-CD95) (Key Resource Table). T-cells were subsequently washed and fixed. All samples were analysed on LSRFortessa (BD Biosciences) and analysed using FlowJo v10.8 (BD Biosciences). Gating was performed as previously described^8^.

### T-cell stimulation and intracellular cytokine staining (ICS) on human PBMC

To investigate the effect of glycaemic variability on the cytokine response of T-cells ex vivo, PBMCs from each donor were stimulated with either i) HKx31 (multiplicity of infection 10), ii) 25ng/mL phorbol myristate acetate and 1µg/mL Ionomycin (PMA/I Sigma Aldrich) iii) CD3/CD28 magnetic beads (Thermo Fisher; Dynabeads) or iii) influenza virus peptide pool (4µg/mL; AnaSpec) (Supplementary Table 1) in RPMI1640 (Gibco) with 10% FCS (Gibco) as described previously^8^.

Stimulations were performed for 18 hours (influenza virus, influenza virus peptide pool) in the presence of anti-CD107a (0.4µg/mL; BioLegend; H4A3), BD GolgiStop and BD GolgiPlug (BD Biosciences). T-cells were subsequently washed and stained for anti-CD3, CD4 and CD8 (Key Resource Table). T-cells were then washed, fixed, and permeabilised using the BD Cytofix/Cytoperm Fixation/Permeabilization kit (BD Biosciences) and stained with anti-MIP1β, anti-IFNγ and anti-TNF (Key Resource Table). Cells were analysed on the LSRFortessa (BD Biosciences) and analysed using FlowJo v10.8 (BD Biosciences). To assess the functionality of these T-cells, the number of MIP-1β+, IFNγ+, TNF+ and CD107a+ cells were assessed according to our previously described gating strategy^8^.

### Statistical analysis

Statistical analyses were performed with GraphPad Prism software (version 9.3.1) (Dotmatics, CA, USA). After determining the normality of distribution with Shapiro-Wilk normality test, outliers within data sets were removed according to the ROUT (1%) test. Continuous data were tested for statistical significance with a Mann-Whitney test or t-test as appropriate. Categorical data were tested for statistical significance with a chi-square test. Multiple linear regression was performed with R software version 4.1.1 (The R Project).

## RESULTS

To investigate the effect of glycaemic variability on anti-viral T-cell populations, 32 participants with T1D were recruited. These participants all had at least two weeks of CGM data available for analysis. Using a CV threshold of 33%, participants were classified as having either low (n = 13) or high (n = 19) glycaemic variability. Participants with low and high glycaemic variability were of equivalent age, sex distribution, type of insulin treatment (injection/pump), body mass index (BMI) and had a similar duration of diabetes (approximately 15 years; Table 1) and recent history of influenza virus infection. Importantly, participants with low and high glycaemic variability had equivalent HbA1c levels and blood glucose levels at the time of sample collection (Table 1). The high glycaemic variability group had a lower rate of influenza virus vaccination within the last 6 months (Table 1). Patients reported taking several other medications asides from those associated with diabetes management, the most common of which was some form of contraception (Supplementary Table 2).

**Table 1:**
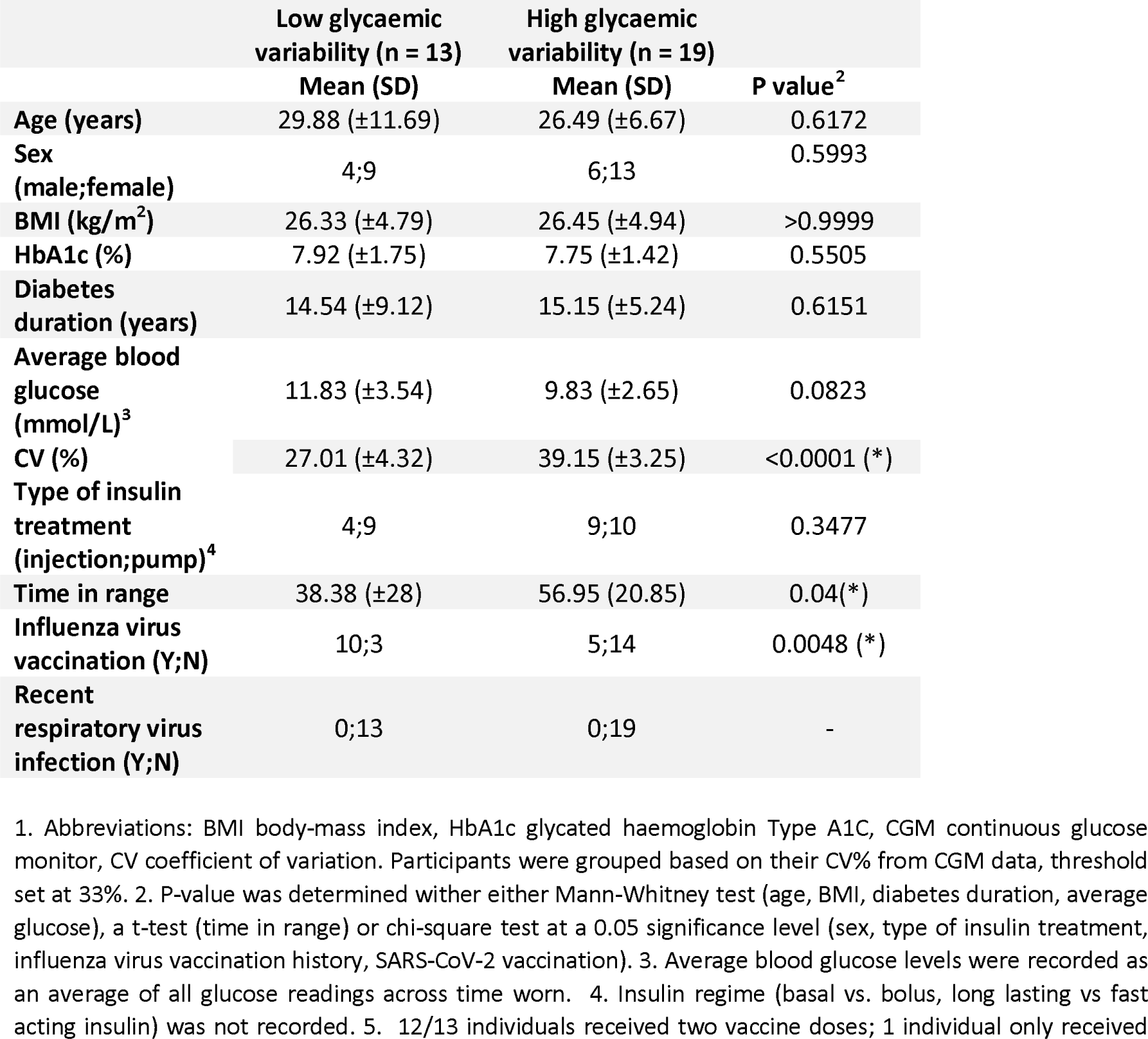
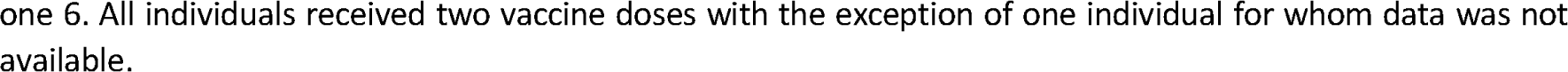
Participant characteristics using a threshold of 33% CV^1^.

|  | Low glycaemic<br>variability (n = 13) | High glycaemic<br>variability (n = 19) | P value<sup>2</sup> |
| --- | --- | --- | --- |
|  | Mean (SD) | Mean (SD) |  |
| Age (years) | 29.88 (±11.69) | 26.49 (±6.67) | 0.6172 |
| Sex<br>(male;female) | 4;9 | 6;13 | 0.5993 |
| BMI (kg/m<sup>2</sup>) | 26.33 (±4.79) | 26.45 (±4.94) | >0.9999 |
| HbA1c (%) | 7.92 (±1.75) | 7.75 (±1.42) | 0.5505 |
| Diabetes<br>duration (years) | 14.54 (±9.12) | 15.15 (±5.24) | 0.6151 |
| Average blood<br>glucose<br>(mmol/L)<sup>3</sup> | 11.83 (±3.54) | 9.83 (±2.65) | 0.0823 |
| CV (%) | 27.01 (±4.32) | 39.15 (±3.25) | <0.0001 (*) |
| Type of insulin<br>treatment<br>(injection;pump)<sup>4</sup> | 4;9 | 9;10 | 0.3477 |
| Time in range | 38.38 (±28) | 56.95 (20.85) | 0.04(*) |
| Influenza virus<br>vaccination (Y;N) | 10;3 | 5;14 | 0.0048 (*) |
| Recent<br>respiratory virus<br>infection (Y;N) | 0;13 | 0;19 | - |

1. Abbreviations: BMI body-mass index, HbA1c glycated haemoglobin Type A1C, CGM continuous glucose monitor, CV coefficient of variation. Participants were grouped based on their CV% from CGM data, threshold set at 33%. 2. P-value was determined with either Mann-Whitney test (age, BMI, diabetes duration, average glucose), a t-test (time in range) or chi-square test at a 0.05 significance level (sex, type of insulin treatment, influenza virus vaccination history, SARS-CoV-2 vaccination). 3. Average blood glucose levels were recorded as an average of all glucose readings across time worn. 4. Insulin regime (basal vs. bolus, long lasting vs fast acting insulin) was not recorded. 5. 12/13 individuals received two vaccine doses; 1 individual only received
one 6. All individuals received two vaccine doses with the exception of one individual for whom data was not available.

To demonstrate the glycaemic profile in high and low glycaemic variability, representative CGM data from each group is shown in Fig. 1A. To evaluate the differences in glycaemic profile between low glycaemic variability and high glycaemic variability with a CV threshold at 33%, the time glucose levels were above and below the healthy range (3.9mmol/L – 10.0mmol/L) were assessed (Fig. 1B). Participants with high glycaemic variability had significantly lesser time spent with hyperglycaemic levels, but significantly higher time spent with hypoglycaemic levels (Figure 1B).

**Figure 1.**
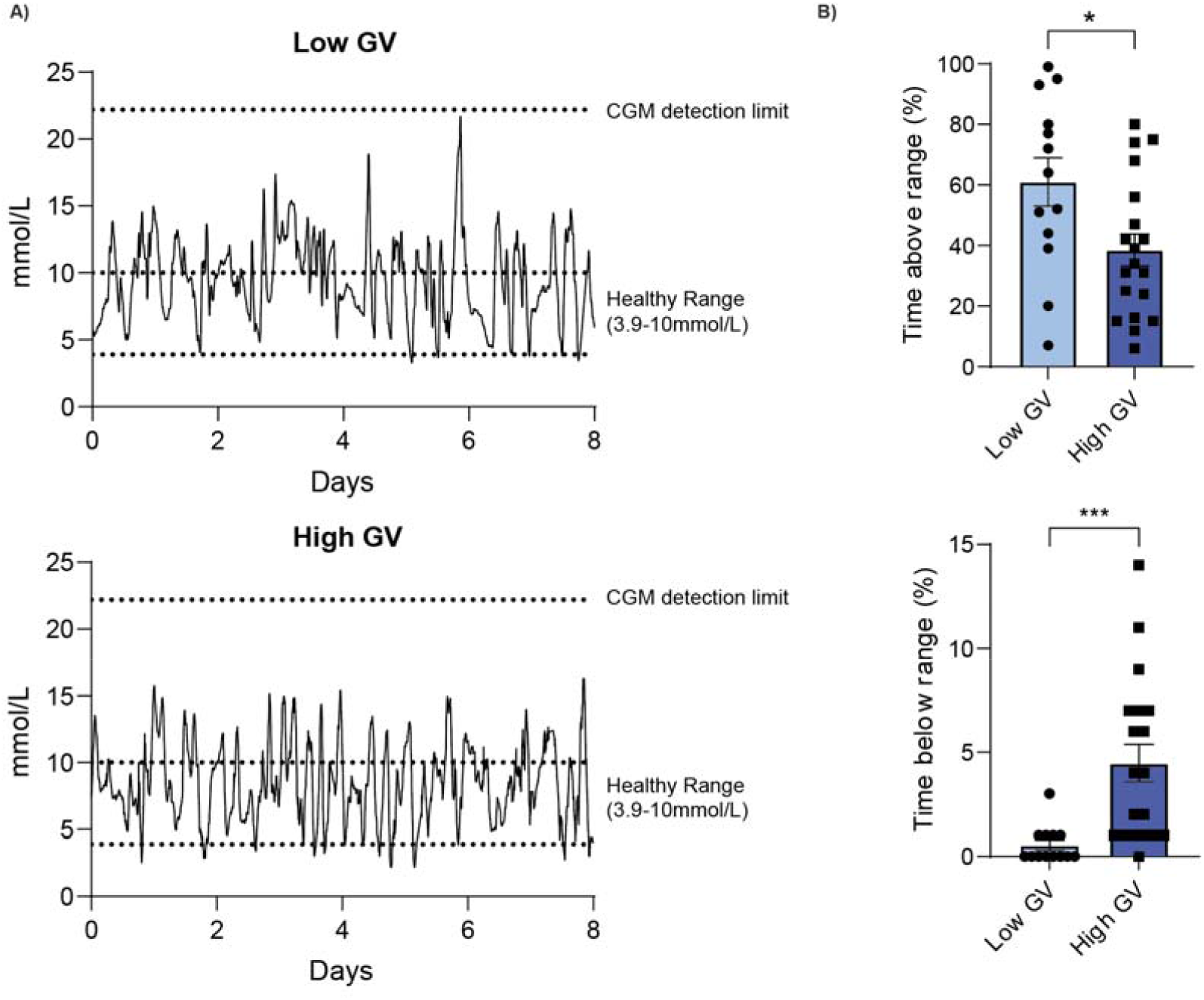
Higher time recorded with hypoglycaemic levels were observed in participants with high glycaemic variability (GV). **(A)**. Representative CGM data for individuals with low glycaemic variability and high glycaemic variability (B). The difference in time glucose levels spent above and below the glycaemic healthy range (3.9mmol/L to 10.0mmol/L) were analysed between both groups. Statistical significance was assessed as described in the Materials and Methods. P< 0.05 *; P<0.001***

To characterise the T-cell populations of both participant groups the percentage of circulating CD4^+^ and CD8^+^ T-cells were assessed. There was no significant difference in the percentage of CD4^+^, CD8^+^, the ratio of CD4:CD8 cells or CD4 CD8 T-cells between those with low glycaemic variability and high glycaemic variability (Fig. 2A). To better define the subsets of CD4 ^+^and CD8^+^ T-cells the percentage of circulating naïve T-cells (T_naive_; CD27^+^CD45RA^+^CD95_-_), stem cell memory T-cells (T_SCM_; CD27^+^CD45RA^+^CD95^+^), central memory T-cells (T_CM_; CD27^+^CD45RA_-_), effector memory T-cells (TEM; CD27^-^CD45RA^-^) and effector memory re-expressing CD45RA T-cells (T_EMRA_; CD27^-^CD45RA^+^) of the total CD4^+^ and CD8^+^ T-cell populations were established (Fig. 2B). No significant differences were seen in the CD4^+^ T-cell subsets between the two participant groups (Fig. 2B). In contrast, patients with high glycaemic variability had a significantly higher percentage of T_naive_ CD8^+^ T-cells but a significantly lower percentage of T_EMRA_ CD8^+^ T-cells compared to patients with low glycaemic variability (Fig. 2B).

**Fig 2.**
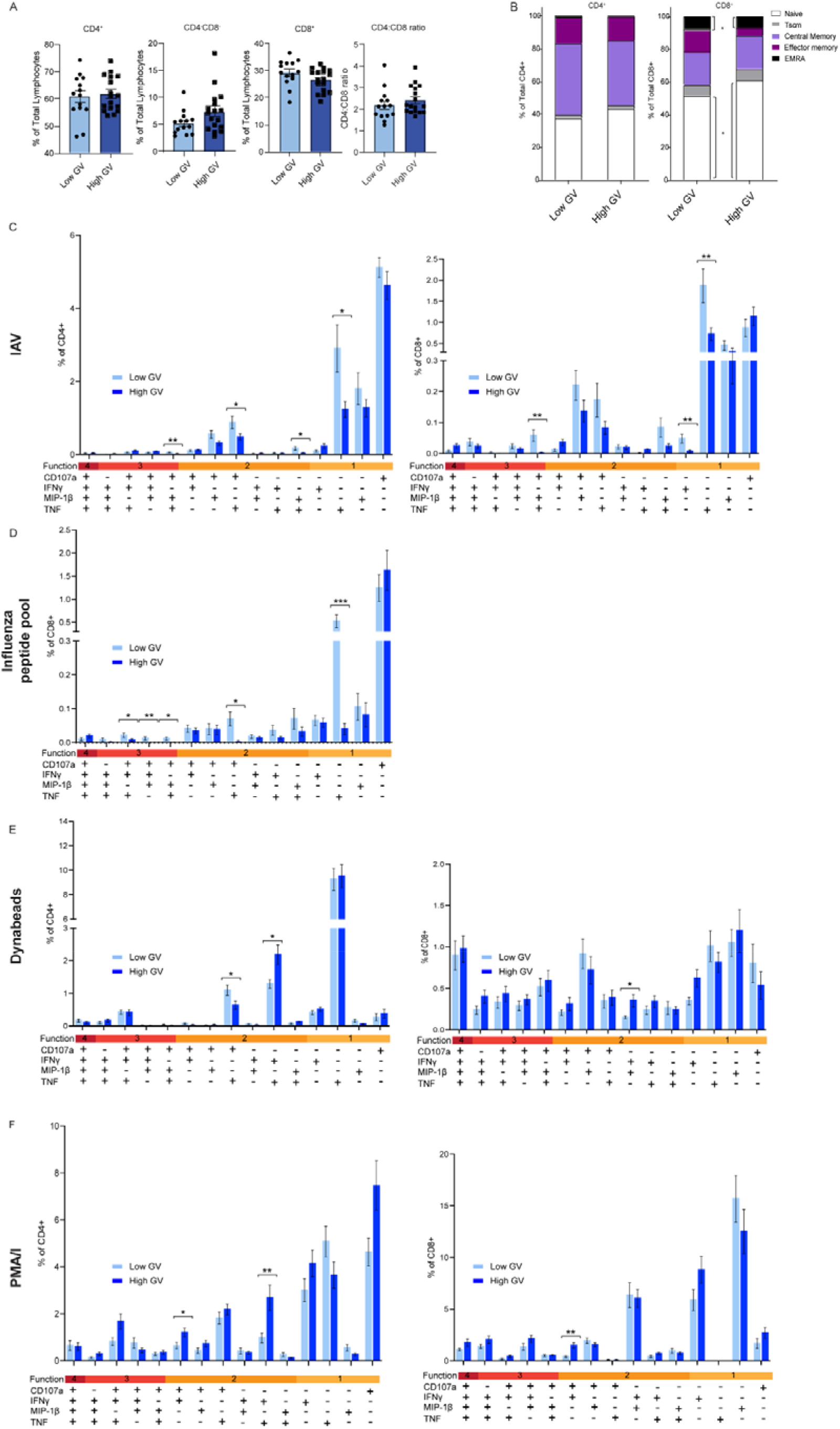
Participants with higher glycaemic variability (GV) exhibit an altered T-cell profile and proportion of TNFα+ cells in response to influenza virus stimulation. PBMC from participants were stained for CD4 and CD8. CD4^+^, CD4^-^CD8^-^ and CD8^+^ positive T-cells (**A**) were stained for CD27, CD45RA and CD95 to further define T-cell subsets (**B**). To study T cell function, Boolean gating analysis was used. PBMC were stimulated with HKx31 (MOI10) (**C**) influenza virus peptide pool (**D**) PMA/I (**E**) and Dynabeads (**F**) for 18 hours. PBMC were then stained for CD107a, IFNγ, MIP-1β and TNF. Statistical significance was determined as described in the Materials and Methods. Bars and error bars represent means (±SEM). Gating was performed as previously described^10^. *: p<0.05.

Next, the cytokine response of T-cells to ex vivo stimulation with infectious influenza virus was assessed (Fig. 2C). In terms of CD4+ T-cells, a lower proportion of CD107a^+^IFNy^-^MIB1b^+^TNF^+^, CD107a^+^IFNy^-^MIB1b^-^TNF^+^ and CD107a^-^IFNy^-^MIB1b^-^TNF^+^ cells were observed in individuals with high GV. In terms of CD8^+^ T-cells, a lower proportion of CD107a^+^IFNy^-^MIB1b^+^TNF^+^, CD107a^-^IFNy^+^MIB1b^-^ TNF^-^ and CD107a^-^IFNy^-^MIB1b^-^TNF^+^ cells were observed in individuals with high GV (Fig. 2C). We then sought to validate these observations in CD8+ T-cells stimulated with an influenza virus peptide pool (Fig. 2D). Consistent with the observations using infectious virus, low glycaemic variability samples treated with influenza virus peptide pool had a significantly higher proportion of CD8^+^CD107a^+^IFNy^-^ MIP1β^+^TNF^+^ relative to healthy controls and high glycaemic variability samples (Fig. 2D). Also consistent with our infectious influenza virus stimulations, high glycaemic variability samples stimulated with influenza peptides had a significantly lower proportion of CD8^+^C107a^-^IFNy^-^MIP1b^-^ TNF^+^ T-cells compared to low glycaemic variability samples (Fig. 2D). When the proportion of any CD8^+^TNF^+^ cell after IAV or IAV peptide pool stimulation was examined significantly higher proportions were still observed in low glycaemic variability samples (Supplementary Fig. 1). Given that a decreased proportion of CD8^+^C107a^-^IFNy^-^MIP1b^-^TNF^+^ cells in high glycaemic variability samples were consistently observed between influenza virus and influenza peptide pool stimulations and showed the largest difference relative to low glycaemic variability samples, we elected to focus on this population.

We next sought to determine if individuals with high glycaemic variability continued to display a decreased proportion of CD8^+^CD107a^-^IFNy^-^MIP1β^-^TNF^+^ cells in response to non-specific stimuli like PMA/I and Dynabeads (CD3/CD28) (Fig. 2E & F). Interestingly, unlike influenza virus stimulations, there were several incidences of CD4+ and CD8+ T-cell populations being higher in individuals with high glycaemic variability following this non-specific stimulation. Furthermore, unlike influenza virus stimulations, there was no significant difference in the proportion of CD8^+^CD107a^-^IFNy^-^MIP1β^-^TNF^+^ between healthy donors, those with high glycaemic variability and those with low glycaemic variability in response to either PMA/I (Fig. 2E) or Dynabeads (Fig. 2F). Taken together, these data suggest that the observed phenotype may be specific to influenza virus or perhaps more broadly viral peptides.

One limitation of these analyses is that some of the observed changes to influenza virus stimulation may be the result of a differential influenza virus vaccination history between those with low and high glycaemic variability (Table 1), although as an inactivated subunit vaccine vaccination does not typically induce strong cellular immunity. Nevertheless, we investigated the ex vivo CD8+ T-cell response follow influenza virus stimulation exclusively in influenza vaccinated individuals. Consistent with our original observations influenza vaccinated individuals with low glycaemic variability had a higher proportion of CD8^+^CD107a^+^IFNy^-^MIP1β^+^TNF^+^ cells (Supplementary Fig. 2). Furthermore, those with low glycaemic variability had a trend towards an increased percentage of CD8^+^ CD107a^-^IFNγ^-^ MIP-1β^-^TNF^+^ T-cells (p = 0.05; Supplementary Fig. 2). This held true when the proportion of any CD8^+^TNF^+^ cell after IAV stimulation was examined (Supplementary Fig. 2). To further complement these results, we performed a multiple linear regression model looking at the relationship between glycaemic variability, influenza vaccination status and the percentage of CD8+CD107a^-^IFNγ^-^MIP-1β^-^ TNF^+^ T-cells. Importantly glycaemic variability (high vs low) had a significant relationship with the percentage of CD8+ CD107a^-^IFNγ^-^MIP-1β^-^TNF^+^ (p = 0.04) T-cells whilst influenza vaccination did not (p = 0.350).

## DISCUSSION

Individuals with diabetes can experience significant intra and inter-day fluctuations in blood glucose levels. To date, the effect of these fluctuations on the T-cell response to influenza virus, or other viral diseases, has been undefined. Here, we provide the show that this glycaemic variability is associated with changes in the T-cell population and reduced CD8^+^ T cell TNF production to ex vivo stimulation with influenza virus.

Patients with high glycaemic variability had a significantly higher percentage of T_naïve_ CD8^+^ T-cells compared to patients with low glycaemic variability. Considering the clinical evidence that high glycaemic variability is associated with more severe viral disease^4,9-13^ it is interesting to speculate what functional consequences increased T_naïve_ CD8^+^ T-cell population may have. Whether this represents a population of T-cells that is unable to differentiate into functional effector or memory cells (which would then likely be associated with an impaired immune response) remains to be determined. Therefore, the clinical consequences of these observations, if any, require further investigation.

In the present study we observed a reduced proportion of CD8+CD107α-IFNγ-MIP1β-TNF+ T-cells in response to stimulation with influenza virus. This same phenomenon was not observed following stimulation with non-specific stimuli (such as PMA/I or Dynabeads). This suggests that high glycaemic variability does not impair CD8+T cell function *per sae* rather more specifically the response to influenza virus or viral stimulation. The production of TNF by CD8+ T cells during a viral infection is essential for the induction of apoptosis, the recruitment and activation of other immune cells, CD8+ T-cell function and the regulation of anti-viral immunity. However, TNF produced by CD8+ T-cells during influenza virus infection can also cause ‘bystander damage’ of pulmonary epithelial cells, resulting in immunopathology^21^. Therefore, whether these observations contribute to the increased influenza severity in individuals with diabetes with high glycaemic variability^4,9-13^ remains to be determined. Nevertheless, these data provide the evidence that assessing glycaemic variability may provide an important insight into the anti-viral CD8+ T-cell response of individuals living with diabetes.

The precise mechanism by which high glycaemic variability may reduce the TNF response of CD8 T-cells and affect the proportion of T_naive_ and T_EMRA_ CD8^+^ T-cells remains unclear. There is a growing body of evidence that glycaemic variability is associated with increased oxidative stress and the production of radical oxygen species (ROS) relative to steady state hyperglycaemia ^22^. This may directly affect T-cell function. For example, short pre-exposure of human PBMC to H_2_O_2_ reduces the ability of activated/memory CD3^+^ and CD8^+^ T-cells (CD45RO^+^) to produce key effector cytokines such as IFNγ and TNF^23^. This effect may be more pronounced in activated/memory T-cells ^23^. Similarly, over production of ROS is known to drive naïve T-cell proliferation^24,25^. Whether these changes in circulating T-cell population were the result of oxidative stress or another biological system affected by glucose fluctuations in the blood remain to be determined.

Our study has some limitations that are important to acknowledge. Firstly, in the participant cohort the sample size was low and glycaemic variability was categorized based on two weeks of CGM data. Whether additional effects would have been detected if a longer period of CGM data were available remains to be determined. It is also important to note that the participants recruited to this study all had T1D. This patient group was selected as CGMs are more common amongst those living with T1D. Interestingly, recent evidence suggests that in patients with T2D who did wear CGMs glycaemic variability was associated with changes in the T cell population (namely an altered Th1/Th2 ratio and frequency of Tregs) ^26^. These data suggest that the differences observed in T cell populations observed herein may not be restricted to patients with T1D^26^. Finally, additional patient information such as GAD positivity or beta cell function was not recorded and may have influenced the results.

In sum, this study has provided evidence that glycaemic variability, rather than steady-state hyperglycaemia, effects T cell responses to influenza virus. These data have important implications for clinical practice. Specifically, these data suggest that glycaemic variability provides a possible predicative approach to assess anti-viral immunity, thus providing better management of infectious diseases in this patient population. Indeed, these data represent a further impetus to make CGMs widely available to patients with both T1D and T2D.

## Supporting information

STAR methods

## Data Availability

All data produced in the present study are available upon reasonable request to the authors

## ACKNOWLEDGEMENTS

We would like to thank all individuals who participated in this study. This project was supported by the National Health and Medical Research Council (NHMRC; APP1159959). KRS is supported by NHMRC investigator grant 2007919. CES was supported by the ARC-DECRA Fellowship (#DE220100185) and the University of Melbourne Establishment Grant. KK was supported by the NHMRC Leadership Investigator Grant (#1173871). EJG is supported by an Australian Research Council (ARC) DECRA Fellowship (DE210101479). SG is supported by an NHMRC Senior Research Fellowship (#1159272). L.C.R. is a recipient of a NHMRC EL1 Fellowship (#2026357).

The authors declare no competing interest.

