## Supplementary Material for "High glycaemic variability in individuals with type 1 diabetes is associated with a reduced proportion of CD8^+^ TNF^+^ cells in response to influenza A virus"

**Supplementary Table 1: influenza virus peptides in influenza peptide pool**

| HLA-I allele | Peptide sequence |
| --- | --- |
| A1 | VSDGGPNLY |
| A1 | CTELKLSDY |
| A2 | GILGFVFTL |
| A2 | FMYSDFHFI |
| A68 | KTGGPIYKR |
| A3 | RVLSFIKGTK |
| A3 | ILRGSVAHK |
| A3, A11, A60B1 | SIIPSGPLK |
| B7 | LPFDKTTVM |
| B8 | ELRSRYWAI |
| B27 | SRYWAIRTR |
| B27 | ASCMGLIY |

**Supplementary Table 2: Patient medication usage (excluding those associated with diabetes management)**

| Low Glycaemic Variability (n = 13) | High Glycaemic variability (n = 19) |
| --- | --- |
| Estelle 35 (1/13)  Ryaltris (1/13)  Rosuvastatin (1/13)  Ramipril (2/13)  Fluoxetine (3/13)  Finasteride (1/13)  Doxycycline (1/13)  Femtab 2100 (1/13)  Ventolin (1/13)  Flixotide (1/13)  Implanon (1/13) | Gabapentin (1/19)  Thyroxine (1/19)  Salbutamol (1/19)  Cefalexin (1/19)  Levelen (1/19)  Atorvastatin (1/19)  Perindopril/indapamide (1/19)  Amlodipine (1/19)  Fluvoxamine (1/19)  Pantoprazole (1/19)  Unspecified contraceptive (1/19) |

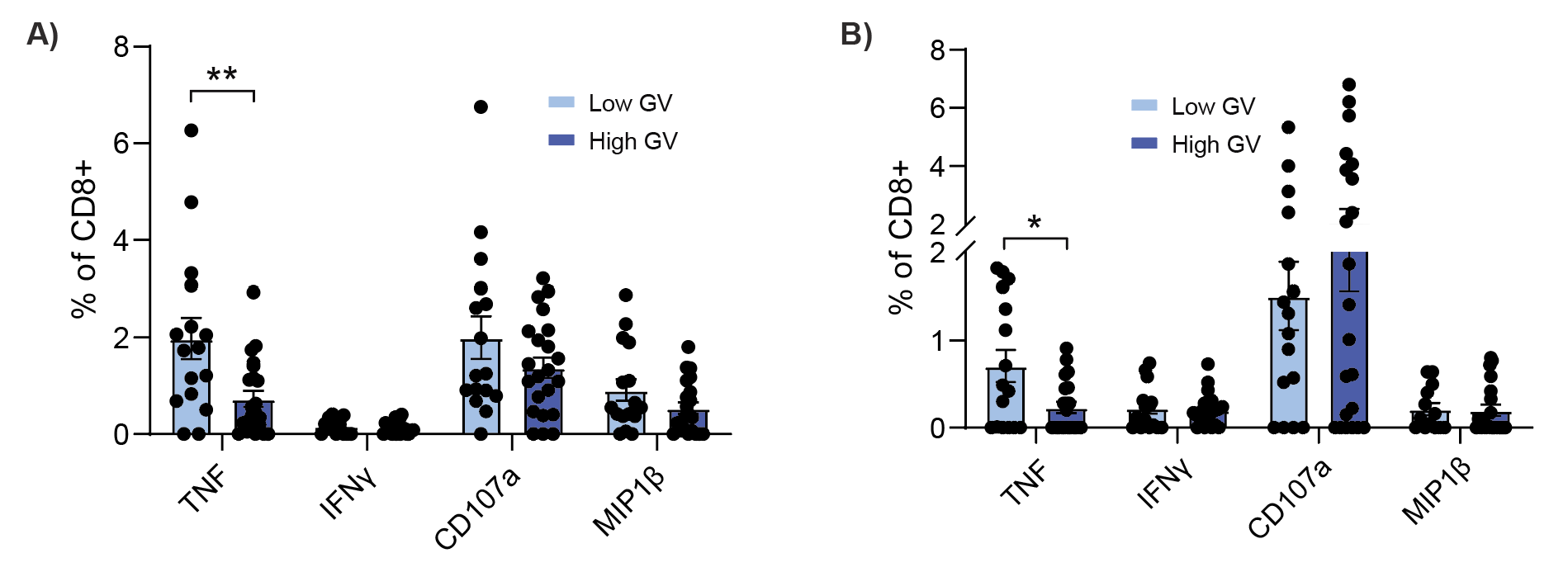

**Supplementary Fig. 1: Participants with higher GV exhibit a reduced percentage of TNF^+^CD8^+^ T-cells.** PBMC from participants living with T1D and who were virus were stimulated with **A)** HKx31 (MOI10) or **B)** influenza peptide pool for 18 hours. PBMC were stained for surface markers CD4 and CD8, and activation markers CD107a, IFNγ, MIP-1β and TNF. Statistical significance was determined as described in the Materials and Methods. Bars and error bars represent means (±SEM). Gating was performed as previously described^10^. *: p<0.05.

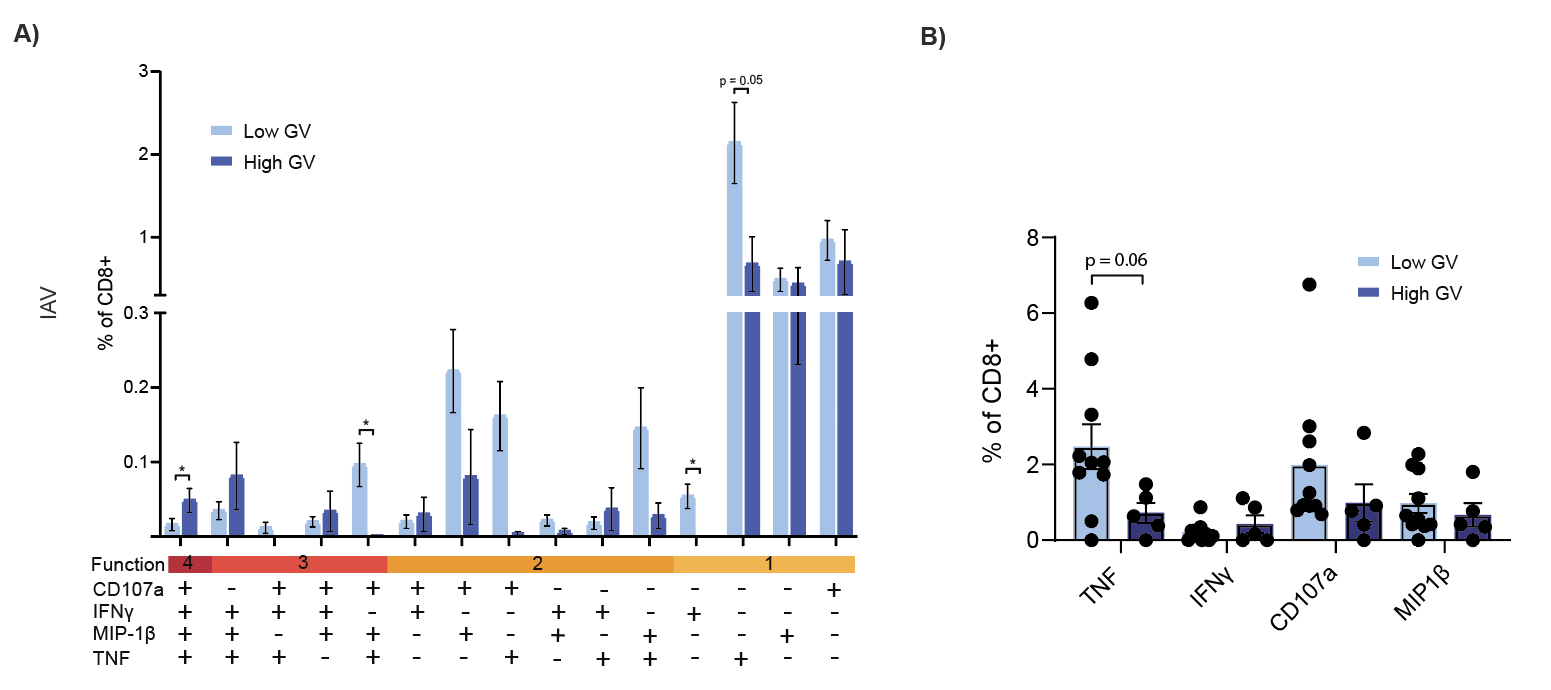

**Supplementary Fig. 2: Participants with higher GV and influenza vaccination history exhibit altered CD8 T-cell derived cytokines.** PBMC from participants living with T1D and who were vaccinated against influenza virus were stimulated with HKx31 (MOI10) for 18 hours. PBMC were stained for surface markers CD4 and CD8, and activation markers CD107a, IFNγ, MIP-1β and TNF. **A)** Subsets of cytokine producing CD8^+^ T-cells. **B)** Percentage of total cytokine CD8^+^ T-cells. Statistical significance was determined as described in the Materials and Methods. Bars and error bars represent means (±SEM). Gating was performed as previously described^10^. *: p<0.05.
