## Supplementary material for "High glycaemic variability in individuals with type 1 diabetes is associated with a reduced proportion of CD8^+^ TNF^+^ cells in response to influenza A virus": STAR methods

### Key Resources table

| REAGENT or RESOURCE | SOURCE | IDENTIFIER |
| --- | --- | --- |
| **Antibodies (PBMC T cell subset characterisation)** | | |
| Anti-CD3 (UCHT1) BV480 | BD Biosciences | Cat #566166; RRID AB_2739563 |
| Anti-CD4 (SK3) BV650 | BD Biosciences | Cat #563875; RRID AB_2744425 |
| Anti-CD8 (SK1) PerCP5.5 | BD Biosciences | Cat #565310; RRID AB_2687497 |
| Anti-CD14 (MØP9) APC-Cy7 | BD Biosciences | Cat #560180; RRID AB_1645464 |
| Anti-CD19 (HIB19) APC-Cy7 | BD Biosciences | Cat #560727; RRID AB_1727437 |
| Anti-CD27 (M-T271) APC | BD Biosciences | Cat #558664; RRID AB_1645457 |
| Anti-CD45RA (HI100) FITC | BD Biosciences | Cat #555488; RRID AB_395879 |
| Anti-CD95 (DX2) BV421 | BD Biosciences | Cat #562616; RRID AB_2737679 |
| Live/Dead stain Near Infrared | ThermoFisher Scientific | Cat #L34976 |
| **Antibodies (PBMC stimulation)** | | |
| Anti-CD3 (UCHT1) PE-Cy7 | BD Biosciences | Cat #563423; RRID AB_2738196 |
| Anti-CD4 (RPA-T4) PE | BD Biosciences | Cat #555347; RRID AB_395752 |
| Anti-CD8 (SK1) PerCP5.5 | BD Biosciences | Cat #565310; RRID AB_2687497 |
| Anti-TNF (Mab11) AF700 | BD Biosciences | Cat #557996; RRID AB_396978 |
| Anti-IFNγ (B27) V450 | BD Biosciences | Cat #560371; RRID AB_1645594 |
| Anti-MIP-1β (D21-135) APC | BD Biosciences | Cat #560686; RRID AB_1727565 |
| Anti-CD107a (H4A3) FITC | BD Biosciences | Cat #560949; RRID AB_396134 |
| Live/Dead stain Near Infrared | ThermoFisher Scientific | Cat #L34976 |
| **PBMC stimulants** | | |
| phorbol myristate acetate | Sigma Aldrich | Cat #P8139 |
| Ionomycin | Sigma Aldrich | Cat #I0634 |
| CD3/CD28 magnetic beads | ThermoFisher Scientific | Cat #11161D |
| influenza virus peptide pool | AnaSpec | Cat #AS-62340 |
| **Critical Commercial assays** | | |
| BD CytoFix/Cytoperm plus fixation/Permeabilization solution Kit with BD GolgiPlug | BD Biosciences | Cat #555028; RRID AB_2869013 |
| **Software and Algorithms** | | |
| Prism 7 | Graphpad | Version 7.0; RRID:SCR_002798 |
| FlowJo V10.8 | FlowJo | Version 10.0; RRID:SCR_008520 |
| R software v4.1.1 | R project | Version 4.1.1; RRID:SCR_001905 |

### Resource availability

#### Lead contact

Please contact Kirsty Short at for resource access.

#### Materials availability

This study did not generate new unique reagents.

#### Data and code availability

All data reported in the paper are available from the lead contact upon request. This paper does not report any original code. Any additional information required to reanalyse the data reported in this paper is available from the lead contact upon request.

### Experimental model and study participant details

**Participant recruitment**

A cohort of 32 people with clinically diagnosed T1D who were routinely using continuous, or flash glucose monitoring (CGM) devices were recruited between 8/6/21 and 11/11/21. Inclusion criteria were 18-60 years of age, not pregnant at the time of study, non-smokers of nicotine cigarettes, minimum diabetes mellitus duration of two years and no known immune disease requiring immunosuppressants. Blood, clinical data, and two weeks of prior CGM data from these participants were collected at the point of recruitment. To further establish the quality of CGM data, only samples with CGM time worn above 70% were included in the analysis. These participants were subsequently grouped based on their coefficient of variation (CV; glucose SD expressed as a percentage of the mean glucose) reading from their CGM with the threshold set at 33.0%. To establish a cohort of individuals without diabetes, 16 participants without any known health conditions were recruited from 1/8/20 to 1/4/21. This study was approved by Mater Research Ethics Committee (HREC/MML/55151 V2; HREC/17/MHS/78) and the University of Queensland Ethics Committee (2019/HE002522). All methods were performed in accordance with institutional guidelines and regulations. Written consent was obtained from all study participants.
